# Informing patterns of health and social care utilisation in Irish older people according to the Clinical Frailty Scale

**DOI:** 10.1101/2021.04.21.21255882

**Authors:** Aisling M. O’Halloran, Peter Hartley, David Moloney, Christine McGarrigle, Rose Anne Kenny, Roman Romero-Ortuno

**Author notes:** Corresponding Authors: Dr Aisling O’Halloran and Prof. Roman Romero-Ortuno, The Irish Longitudinal Study on Ageing (TILDA), Trinity Central, 152-160 Pearse Street, Dublin 2, D02 R590, Ireland. and. Funding: TILDA is funded by Atlantic Philanthropies, the Irish Department of Health and Irish Life. RRO is funded by a Grant from Science Foundation Ireland under Grant number 18/FRL/6188. The funders had no role in the conduct of the research and/or preparation of the article; in study design; in the collection, analysis and interpretation of data; in writing of the report; or in the decision to submit the paper for publication.

## Abstract

There is increasing policy interest in the consideration of frailty measures (rather than chronological age alone) to inform a more equitable allocation of health and social care resources in the community. The Clinical Frailty Scale (CFS) has attracted interest for its simplicity and consideration of multiple relevant geriatric dimensions. However, a criticism of the CFS has been the possible subjectivity in the scoring, bringing the possible danger of lack of scoring consistency across agencies. For that reason, the authors of the CFS published a classification tree method to assist with routine scoring of the CFS.

The aim of the present study was to apply the CFS classification tree to data from adults aged 65 and over from The Irish Longitudinal Study on Ageing (TILDA) and correlate derived CFS categories with patterns of health and social care utilisation in Irish older people assessed in Wave 5 of the study (year 2018). In addition, we explored how CFS categories and states changed over 8 years in TILDA between Wave 1 (2010) and Wave 5.

Results showed the following prevalence of CFS categories in Wave 5: 6% ‘very fit’ (CFS1), 36% ‘fit’ (CFS2), 31% ‘managing well’ (CFS3), 16% ‘vulnerable’ (CFS4), 6% ‘mildly frail’ (CFS5), 4% ‘moderately frail’ (CFS6) and 1% ‘severely frail’ (CFS7). No participants were ‘very severely frail’ or ‘terminally ill’. In wave 5, increasing CFS categories had an association with increasing utilisation of hospital and community health services, and increasing hours of formal and informal social care provision. The transitions analyses from Wave 1 to 5 suggested a dynamic picture of CFS transitions, with 2-year probability of transitioning from ‘fit’ (CFS1-3) to ‘vulnerable’ (CFS4), and ‘fit’ to ‘frail’ (CFS5+) at 34% and 6%, respectively. ‘Vulnerable’ and ‘frail’ had a 22% and 17% probability of reversal to ‘fit’ and ‘vulnerable’, respectively.

Our results suggest that the CFS classification tree was able to stratify the TILDA population aged 65 and over into subgroups with increasing health and social care needs. The CFS classification tree could be used to aid the allocation of health and social care resources in older people in Ireland, but given the frequency of CFS transitions in the population, it is recommended that CFS status in individuals is reviewed at least every 2 years.

## Introduction

Frailty is a common condition in older adults, although it is not an inevitable part of ageing (1). Frailty occurs when multiple body systems have gradually lost their inbuilt reserves and can occur in any older adult (i.e. aged 65 and over), but it becomes more prevalent at more advancing ages (2). Frailty is not a medical diagnosis because it can have different drivers in different individuals. Yet, frailty represents a state of vulnerability in the older person that has been consistently associated with premature loss of independence and adverse health outcomes, independently of chronological age (3, 4).

Older adults living with frailty are at an increased risk of sudden deterioration in their health following exposure to insults that robust people can more easily withstand. This has become very evident during the COVID-19 pandemic and its aftermath (5, 6). Not only has frailty been associated with increased risk of COVID-19 mortality in older adults (7), but in survivors and ‘post-cocooning’ individuals, an increased incidence of deconditioning and need for increased rehabilitative and social care supports have been evident for individuals, families and clinicians alike (8).

Frailty is intended to capture the usual ‘baseline’ of an older individual, i.e. in the absence of any acute illness that may be confounding the assessment. The frailty paradigm predicts that when a frailer older individual survives an acute illness, there is a higher risk of functional loss, but this will often recover to the pre-existing baseline after a period of rehabilitation. However, in a minority of cases, there may be a permanent functional loss requiring new supports (2). This is what clinicians call ‘a new baseline’. In older adults who survive an acute hospital admission, the risk of acquiring a lower baseline on discharge is estimated at 30% (9). In the community, quantification of this risk has been more difficult due to the lack of routinely collected data. Because having acquired a lower ‘new baseline’ may compromise the individual’s independence, it is important to try to restore baseline function as much as possible both during and after the onset of the acute illness. This often requires access to multidisciplinary rehabilitation resources and social supports. Active rehabilitation during and after the acute illness may reduce the risk of a new baseline occurring, which will require new permanent supports (10).

Even in the absence of known interim acute medical events, frailty is a dynamic process that changes over time and can be viewed on a continuum (11). An older person can transition in either direction between the different states of frailty, namely robustness or non-frailty, pre-frailty (an intermediate sub-clinical state) and frailty (12). Therefore, the regular and proactive identification of people living with frailty in the community could provide an opportunity to develop more effective and equitable healthcare service planning and delivery for older people (13). For example, a ‘dip’ in known frailty status could be a focus for prioritisation of referrals towards the still limited number of integrated care ‘hubs’ (14) that deliver medical assessment and multidisciplinary rehabilitation and ensure that any reversible new disability can be addressed. Indeed, the gold standard for the assessment and management of frailty is Comprehensive Geriatric Assessment (CGA). CGA is a holistic and interdisciplinary assessment of an individual and has been demonstrated to reduce adverse outcomes including disability, cognitive decline, need for long-term residential care and death (10, 15). Even when a lower new baseline has been established despite rehabilitative attempts, frailty identification at that point could also have implications for the prioritisation of social care resources.

It must be noted that even though the identification of frailty may have advantages from the point of view of medical risk stratification and planning of healthcare delivery, the public’s perceptions of frailty are generally negative and many older people with multimorbidity and disability do not identify themselves as frail (16). This is often the case because of a mistaken general perception that ‘nothing can be done’ about frailty, but in fact the correct reading of frailty identification is to facilitate prioritisation and increase equity of access to health and social care resources, where frailty can be fully assessed, addressed and managed.

Despite a lack of agreement on an internationally accepted and easily administered consensus measure of frailty, several methods of screening are commonly used (17, 18). Among the many available tools, the Clinical Frailty Scale (CFS) has attracted interest for its simplicity and consideration of multiple relevant geriatric dimensions, including multimorbidity and degree of symptom control, mobility, physical activity, help with activities of daily living, dependency, and cognition. The CFS does not assess other important geriatric domains (Figure 1), but the assessed dimensions are reasonably comprehensive and can not only be scored by doctors but a wide range of healthcare and allied health professionals (19). In addition, even though the CFS does not directly assess some important dimensions, it can do so indirectly, since geriatric syndromes tend to overlap (Figure 1).

**Figure 1.**
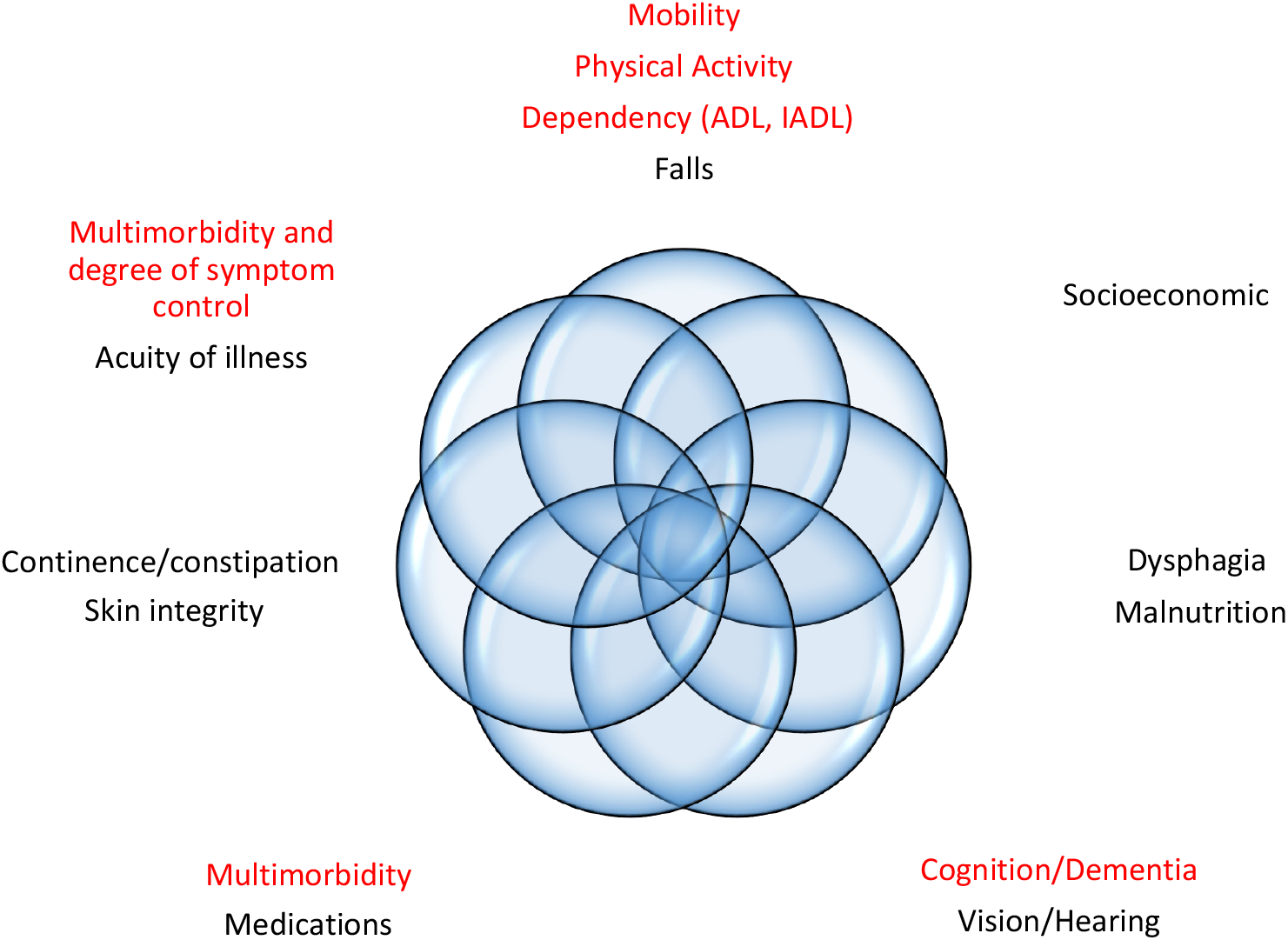
Geriatric dimensions assessed by the Clinical Frailty Scale (in red). ADL: basic activities of daily living (e.g. toileting, transferring, eating). IADL: independent activities of daily living (e.g. managing finances, going shopping, using public transport).

The initial version of the CFS was validated in people aged 65 and older from the Canadian Study of Health and Aging (20) and has been shown to predict mortality, adverse health outcomes and healthcare utilisation in older people across settings (19). Importantly, the CFS is a baseline frailty instrument and even when used in the acute hospital setting, it is intended to capture the status of the individual two weeks before the onset of the acute illness that lead to hospital admission (21). Despite the rolling out of dedicated training modules for more accurate CFS scoring (22, 23), a criticism of the CFS has been the possible subjectivity in the scoring, bringing the possible danger of lack of scoring consistency between individuals and across agencies. However, to aid those concerns, the authors of the CFS have published a classification tree method to assist with routine scoring of the CFS (24). This also allows the retrospective implementation of the CFS in a dataset when the required CFS decision tree variables have been collected.

The CFS classification tree can be implemented to data already collected in The Irish Longitudinal Study on Ageing (TILDA). As such, we saw the opportunity to apply the new CFS classification tree method to the TILDA data and correlated derived CFS categories with patterns of health and social care utilisation in Irish older people. With this, we intended to inform the Irish public and policy makers as to the average levels of health and social care utilisation that can be expected for each of the CFS tree categories. The information presented in this section is based on cross-sectional analyses of TILDA participants from Wave 5 and refers to adults aged ≥65 years living in Ireland in 2018. In addition, following recently developed methodology (12), this report also presents evidence on longitudinal transitions of the CFS in TILDA, to demonstrate the dynamic nature of CFS states over time that will require regular review in populations if any health or social care policies are going to be taking the CFS into account.

## Methods

### Sample

Wave 1 of TILDA (baseline) took place between October 2009 and February 2011, and subsequent data was collected approximately biannually over four longitudinal waves (Wave 2: February 2012 to March 2013; Wave 3: March 2014 to October 2015; Wave 4: January to December 2016; Wave 5: January to December 2018). An overview of the study is available on https://tilda.tcd.ie/about/where-are-we-now/. The full cohort profile is described elsewhere (25).

### Operationalisation of the CFS in TILDA

The CFS was operationalised at waves 1-5 of TILDA according to the published CFS Decision Tree (24). Five items were used in the decision tree to distinguish between CFS classes as follows: (i) number of basic activities of daily living requiring help (BADL); (ii) number of instrumental activities of daily living requiring help (IADL); number of chronic conditions (28 chronic and cardiovascular conditions shown in Appendix 1); (iii) Self-rated health – Excellent, very good, good, fair or poor; (iv) One item from the Center for Epidemiological Studies Depression scale: during the past week how often have you felt that everything that you did was an effort – rarely or none of the time, some or a little of the time, occasionally or a moderate amount of time, all of the time; and (v) moderate or vigorous activity on ≥1 days in the past week as assessed using the International Physical Activity Questionnaire Short Form (IPAQ-SF).

### Cross-sectional analyses of CFS and health and social care service use at Wave 5

For the cross-sectional analyses of CFS and health and social care service utilisation, we employed the most recently completed pre-pandemic wave of TILDA data collection. These data were collected through the computer-assisted personal interview (CAPI) between 16th January 2018 and 1st January 2019.

Descriptive statistics were computed with STATA version 15 (StataCorp, College Station, TX) and given as mean with standard deviation (SD) and range or proportion (%). As described elsewhere (26), we used attrition weights to make estimates representative of the general population aged ≥65 years in Ireland. We examined the prevalence of CFS categories and the association between the CFS categories and utilisation of medical and community-based allied healthcare services. We also examined the levels of informal care and formal community support services that support ageing in place, including hours of paid and unpaid care received per month.

### Longitudinal analyses: CFS transitions over 8 years (Waves 1-5)

The CFS transitions analyses followed the same methodology as a previously published TILDA paper on frailty phenotype transitions (12). The baseline analytical sample included participants who had complete CFS information at Wave 1. For subsequent waves, information was collected on transitions in CFS states and attrition due to deaths or missing data. Mortality was ascertained for all study participants at each follow-up wave. TILDA has approval from Ireland’s General Register Office (GRO) to link survey respondents to their death certificate information held centrally by the GRO, where every death in the Republic of Ireland must be registered (27). Other than deaths, attrition at each wave was classified as ‘missing’.

For the visualisation of the longitudinal CFS transitions, alluvial charts were created using the R ggalluvial package (12). As well as transitions in individual CFS categories, we visualised the transitions in the following CFS states: ‘fit’ (CFS 1-3), ‘vulnerable’ (CFS4) and ‘frail’ (CFS5 or more). In an alluvial plot, the height of the stacked bars at each wave (which represent whether participants’ status for the given frailty state was yes, no, missing or died) is proportional to the number of participants identified as belonging to this state at each wave. The thickness of the streams connecting the stacked bars between waves are proportional to the number of participants who have the state identified by both ends of the stream. To estimate transition probabilities for the CFS states, we used multi-state Markov models using the R msm package, which allows a general multi-state model to be fitted to longitudinal data (12). We obtained matrices of estimated transition probabilities from wave x to wave x + 1 (with 95% confidence intervals [CIs]) for each CFS state.

### Ethical approval

Ethical approval for each wave was obtained from the Faculty of Health Sciences Research Ethics Committee at Trinity College Dublin, Ireland. All participants provided written informed consent prior to inclusion in the study.

## Results

### Sample descriptives: cross-sectional analyses at Wave 5

Of the 8,504 participants recruited to TILDA at Wave 1, 3,279 did not participate in Wave 5, leaving a sample of n=5,225. We removed from our analysis any participant aged less than 65 years of age (n=1,467) and those participants who were not present at Wave 1 (n=108) or who did not have CFS data (n=14). Thus, the analytical sample included n=3,636 participants aged ≥65 years at Wave 5. The average age was 74.5 years, the age range was 65–103 years, and 54.7% were female.

### Sample descriptives: CFS transitions at Waves 1-5

TILDA recruited a total of 8,504 participants at Wave 1, of whom 4,998 (58.8%) were aged less than 65 years. Of the remaining 3506 Wave 1 participants, n=3,503 had complete CFS information. The mean (SD; minimum-maximum) age and sex percentage of participants remaining at the study at each wave were as follows: For wave 1 participants (n=3,503) 73.3 (6.4; 65-105) years and 52.5% female; for Wave 2 (n=2,893): 75.0 (6.2; 67-97) years and 52.6% female; for Wave 3 (n=2,471): 76.9 (5.9; 69-98) years and 53.1% female; for Wave 4 (n=2,096): 78.5 (5.6; 71-101) years and 53.0% female; and for Wave 5 (n=1,743): 80.0 (5.3; 73-103) years and 53.4% female.

### Prevalence of CFS categories at Wave 5

The CFS categories identified in TILDA were: CFS1 - Very fit; CFS2 – Fit; CFS3 - Managing well; CFS4 – Very mild frailty (or vulnerable); CFS5 – Mild frailty; CFS6 - Moderate frailty; and CFS7 - Severe frailty. CFS classes 8 (very severely frail) and 9 (terminally ill) were not observed in TILDA. The prevalence, or the proportion of the community-dwelling population aged ≥65 years, by CFS frailty status at Wave 5, is provided in Figure 2. Results show that 73.6% of adults aged 65 and over were classified as very fit, fit or managing well, while 16% were considered vulnerable or living with very mild frailty. The remaining 10.5% were classified as having some degree of frailty; within the latter, mild frailty was most prevalent (5.5%), followed by moderate (4.3%) and severe frailty (0.7%).

**Figure 2.**
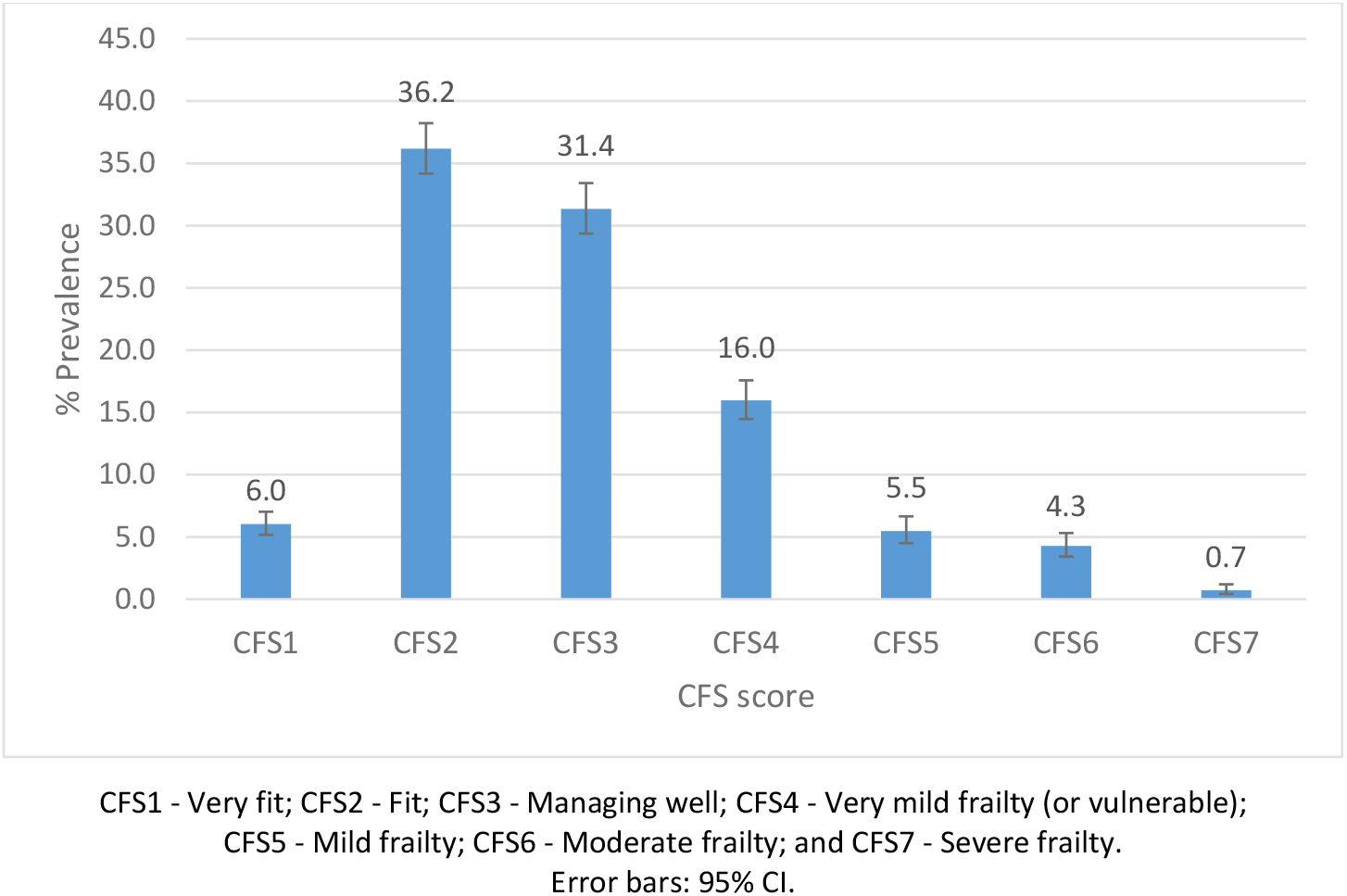
Prevalence of the CFS categories in TILDA Wave 5 (≥65 years)

### Utilisation of medical services and the CFS

At Wave 5, TILDA participants were asked about the number of times they visited a range of medical services including the General Practitioner (GP), a hospital outpatient clinic and the Emergency Department (ED), and the number of overnight hospital admissions, the number of day case procedures and the number of nights spent in hospital over the previous 12 months. We examined the average number of visits in the previous 12 months to each service by older adults in Ireland.

Figures 3, 4 and 5 show the mean number of GP, outpatient and ED visits in the previous twelve months. For each of these services, the mean number of visits increased with higher levels of CFS, for GP (CFS1-CFS7: 2.5-5.9), outpatient (0.7-3.9) and ED (0.1-0.9) visits. This represents an approximately 2-fold, 6-fold and 9-fold increase in utilisation of GP, outpatient and ED services, respectively, between those in the lowest and highest CFS groups.

**Figure 3.**
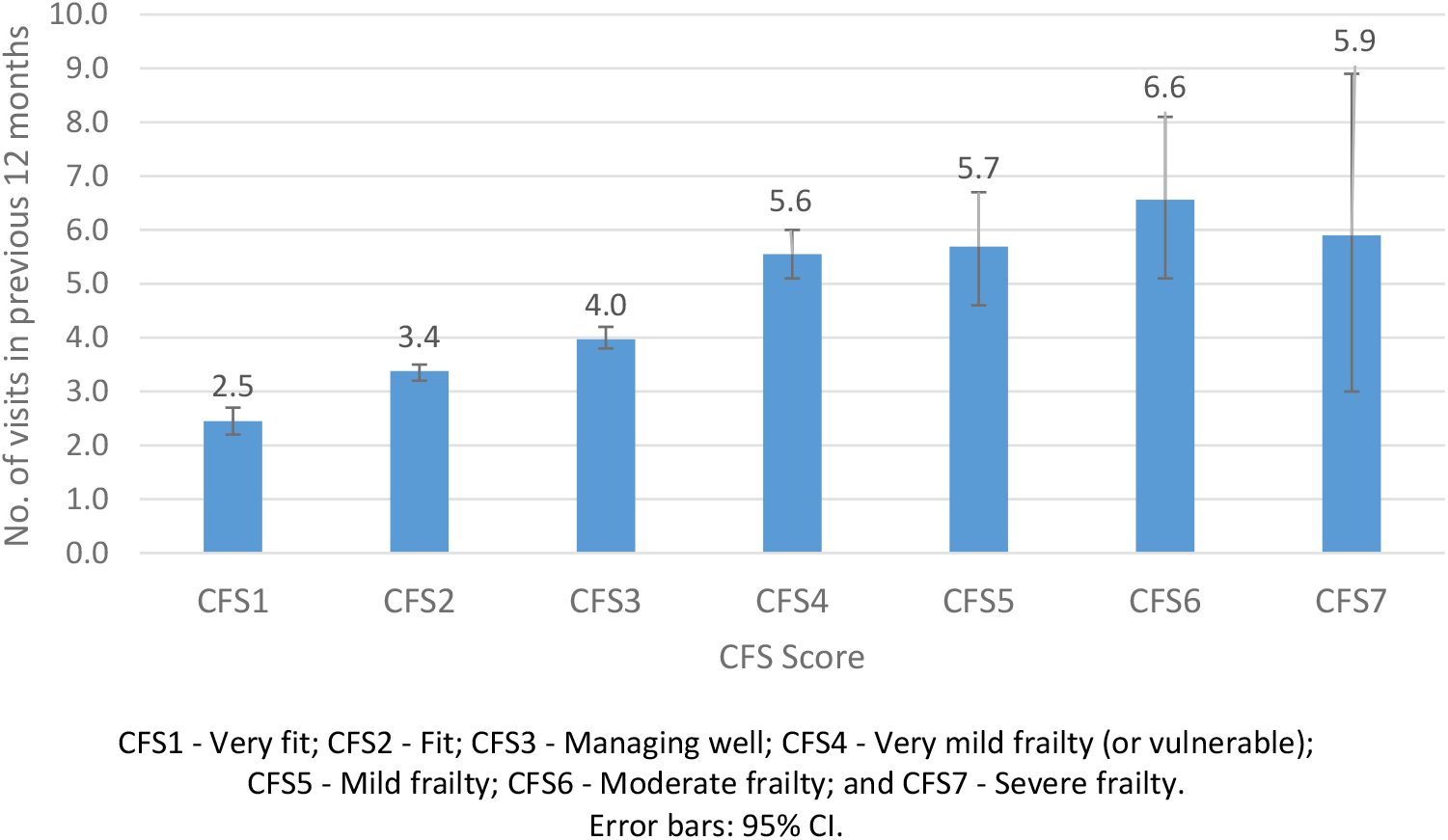
GP visits (mean) in the previous 12 months by CFS

**Figure 4.**
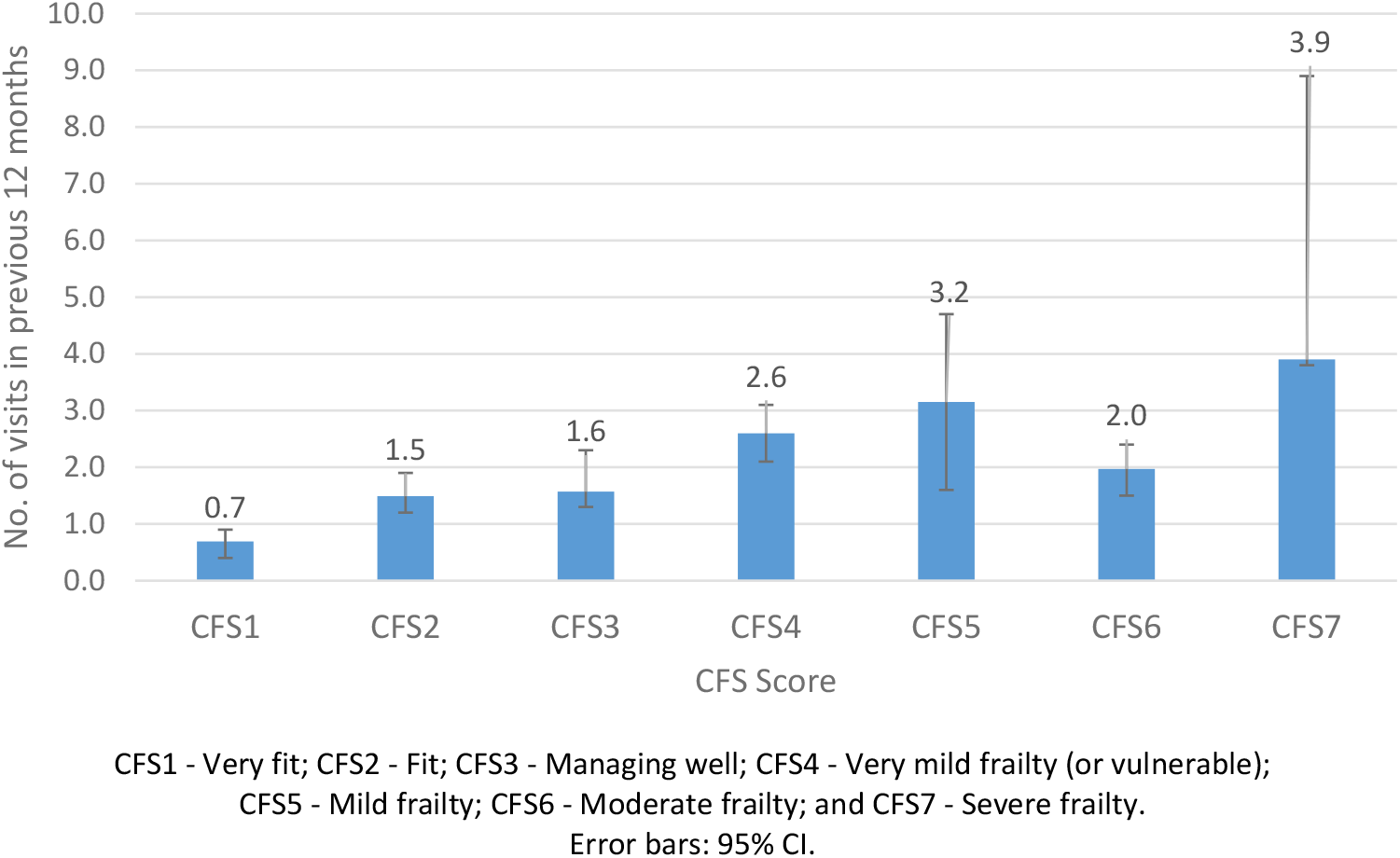
Outpatient clinic visits (mean) in the previous 12 months by CFS

**Figure 5.**
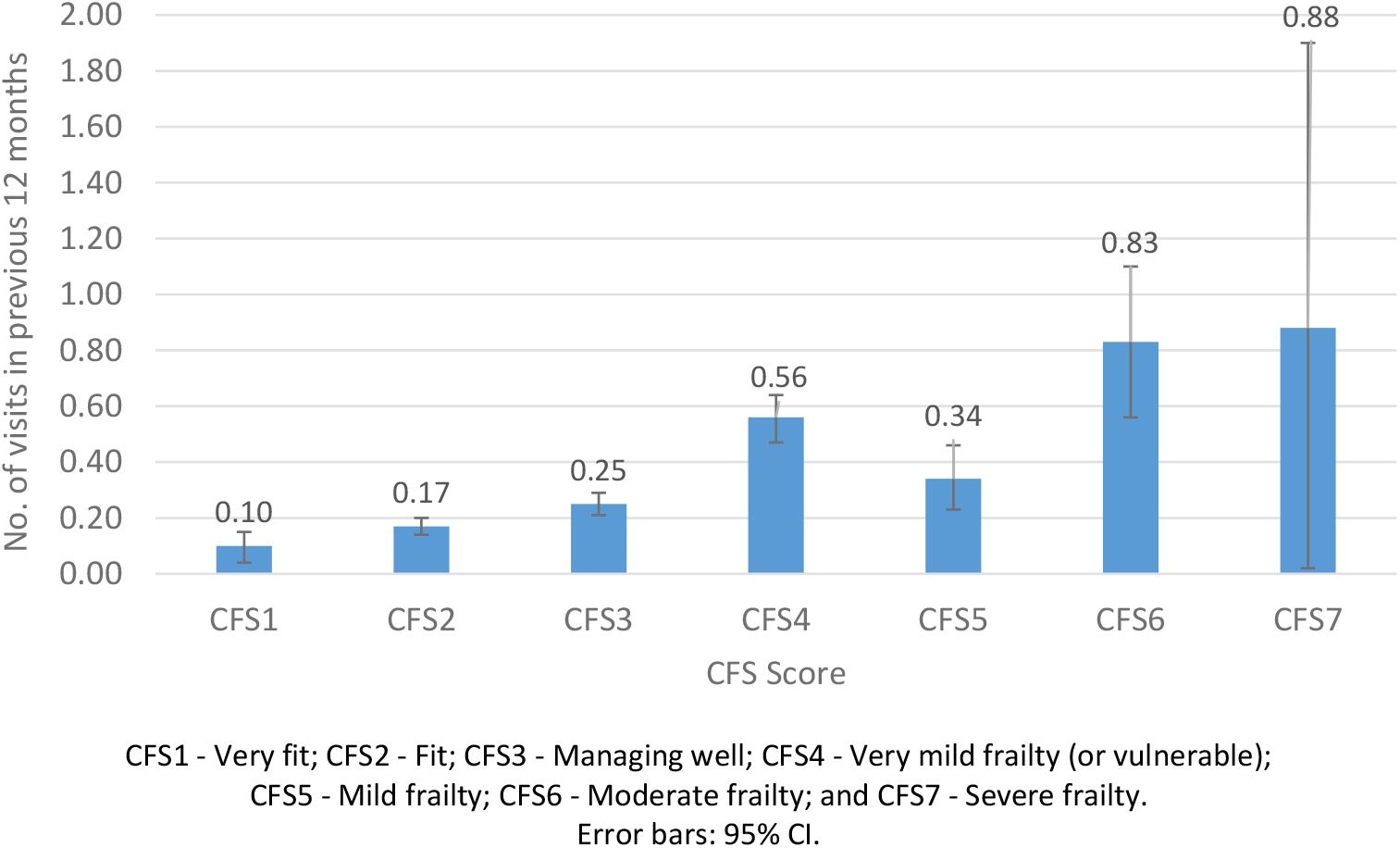
Emergency Department visits (mean) in the previous 12 months by CFS

Figures 6 and 7 show the mean number of overnight hospital admissions and the length of hospital stay (in nights) in the previous twelve months. A similar pattern was observed for overnight hospital admissions (0.2-1.0 admissions) and for length of hospital stay (6.6-26.1 nights), representing a 5-fold and 4-fold increase, respectively, between those in the lowest and highest CFS groups.

**Figure 6.**
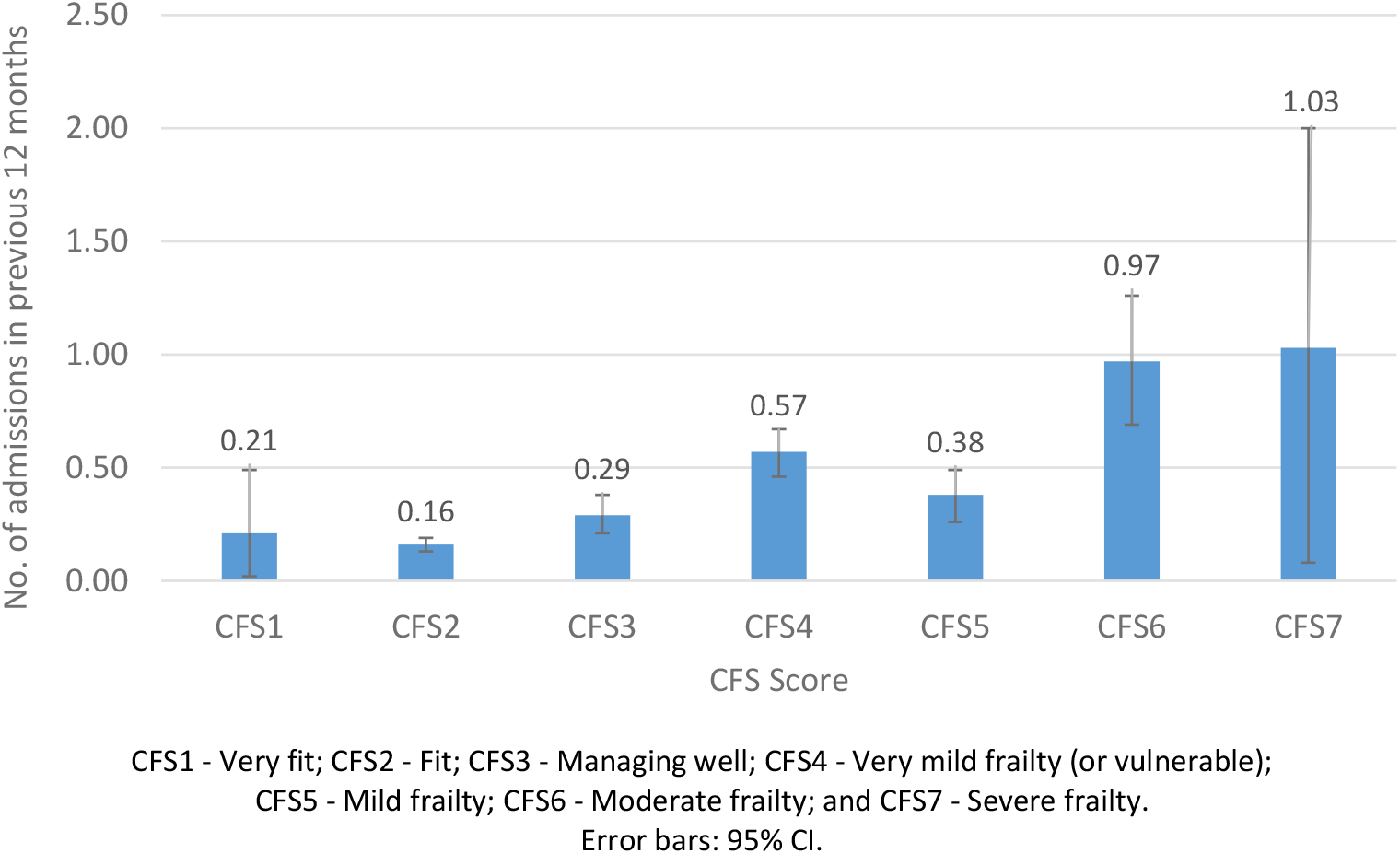
Overnight hospital admissions (mean) in the previous 12 months by CFS

**Figure 7.**
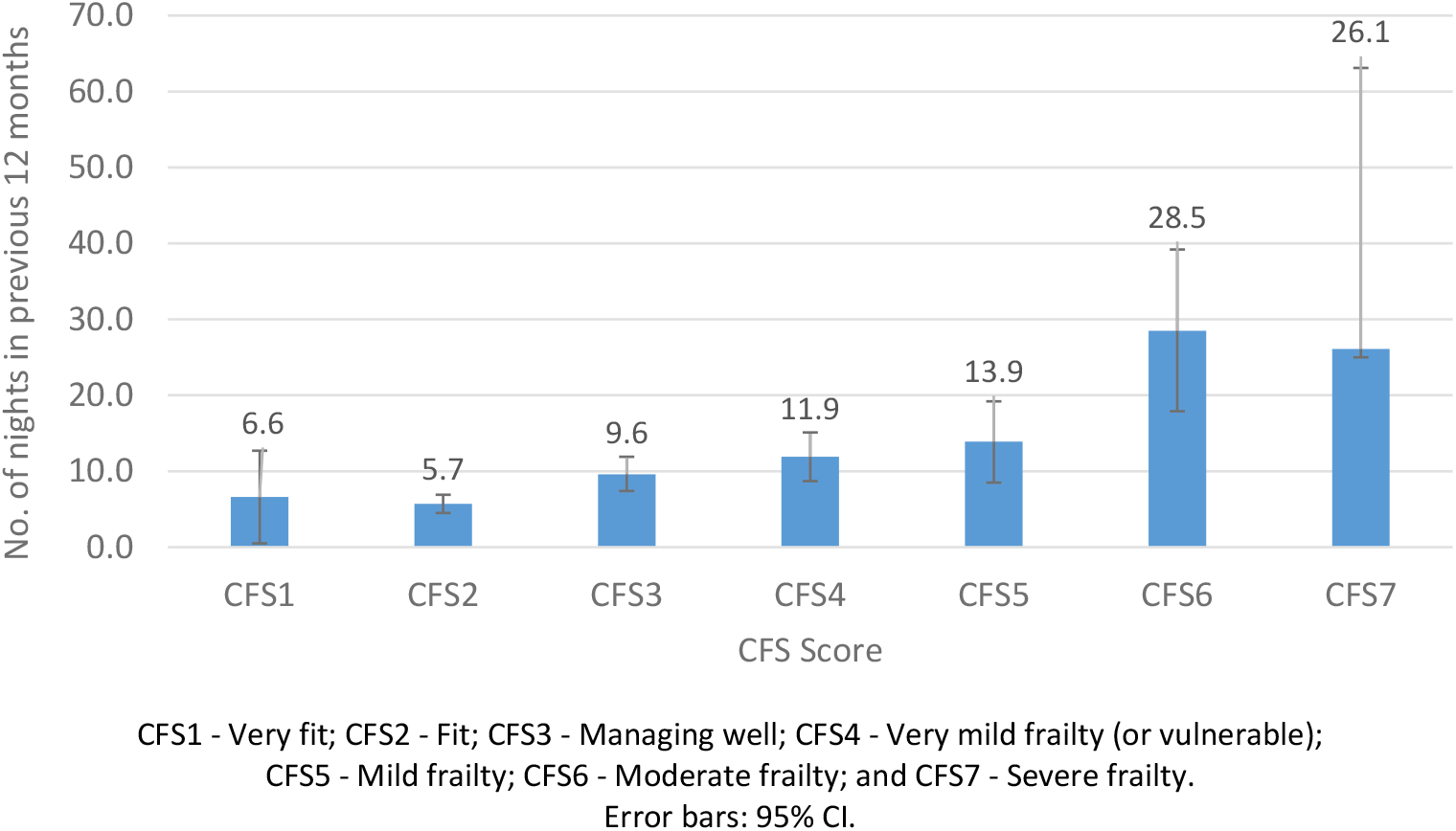
Length of hospital stay (mean nights) in the previous 12 months by CFS

### Utilisation of community-based allied health care and the CFS

In TILDA, data are also collected on community-based healthcare services other than those provided by GPs and hospitals. Participants were asked (yes/no) if they had utilised any of the community-based allied healthcare services in the preceding twelve months, excluding any services for which they had paid anything other than a token or nominal amount. This included state-provided physiotherapy, occupational therapy (OT), public health nurse (PHN), dietician, hearing, dental, optician, psychological/counselling, social work services, speech and language therapy, chiropody, day centre and respite services. Using the binary variable for each of the thirteen services, we generated a count of the mean number of total community services received.

The mean number of state-provided community-based services received and the proportion of each individual service received by CFS groups is provided in figures 8 and 9, respectively. The mean number of state community-based services received increased progressively with higher levels of CFS, from 0.3 for participants classified as CFS1 to 2.0 services for those classified as CFS7.

**Figure 8.**
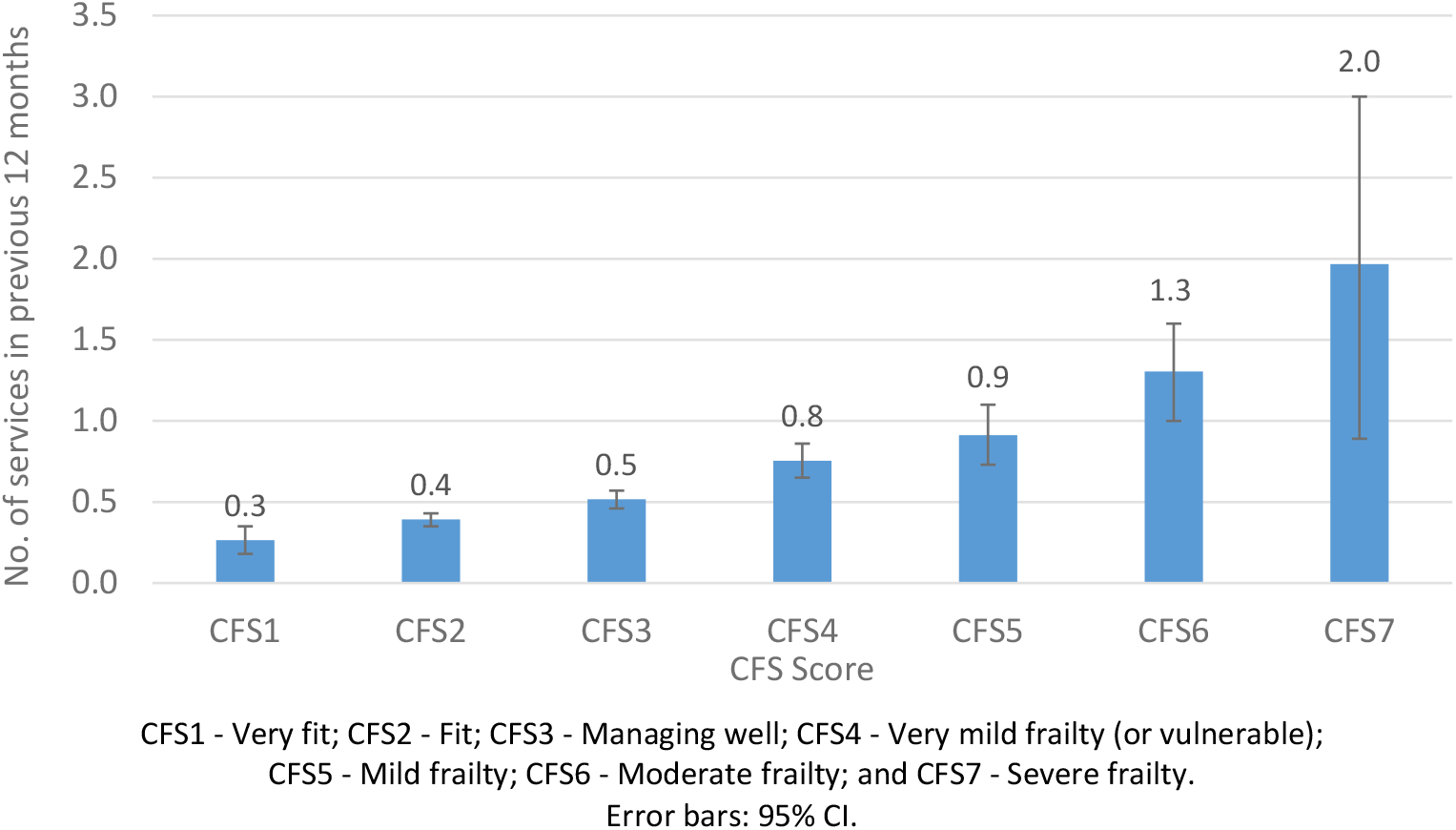
Number of state-provided community-based allied health care services received (mean) in the previous 12 months, by CFS

**Figure 9.**
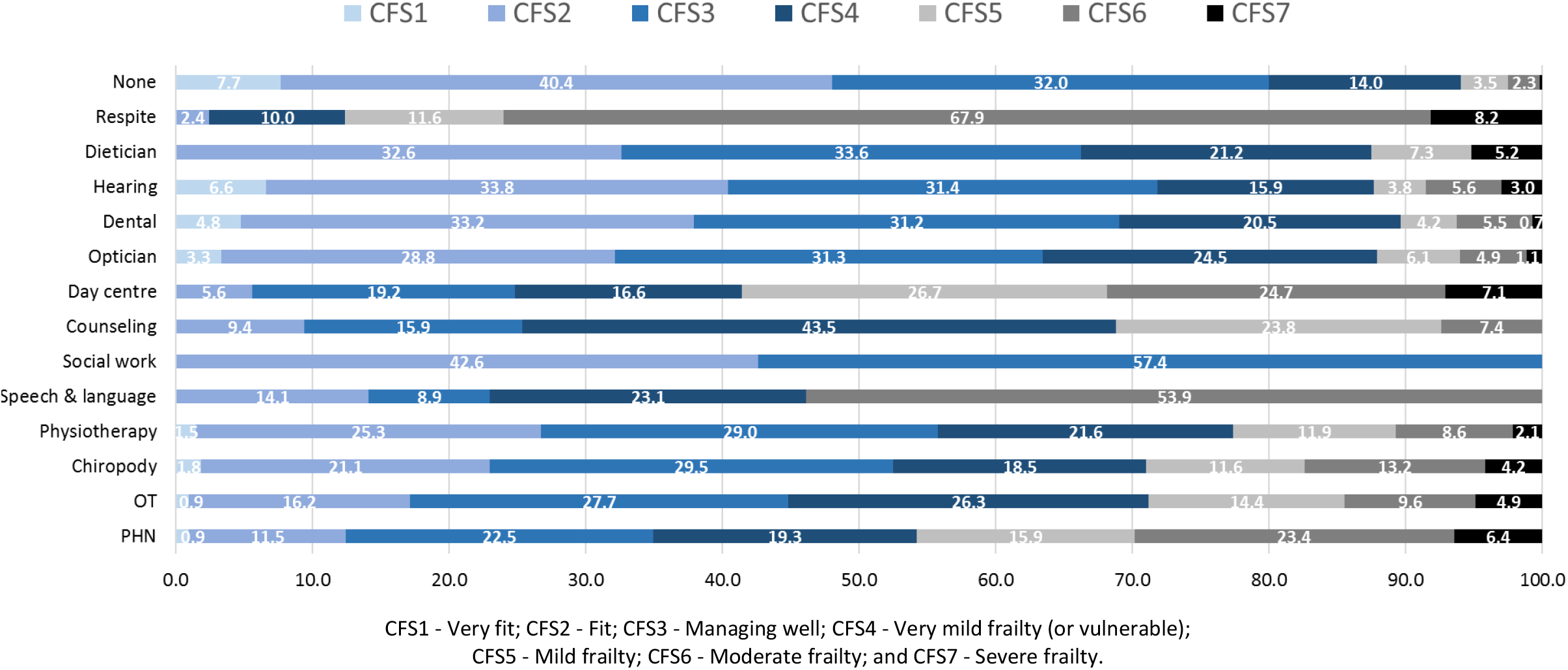
Proportion of people using community-based, public allied health care in the previous 12 months, by CFS

Examining the proportion of each of the thirteen community-based services received by CFS group, there seemed to be a large degree of heterogeneity across the different services (Figure 9). The highest proportions of services received by those classified as having mild to severe frailty (CFS5-CFS7) were: respite care (88%), day centre (59%), speech and language (54%), public health nurse (46%), psychological/counselling (31%), OT and chiropody (30%) and physiotherapy (23%). 13% or less of dental, dietician, optical and hearing/audiology services were received by those living with mild to severe frailty.

### Utilisation of community-based services supporting ageing in place and the CFS

Here we present data on informal care and formal state provided care service, which are delivered in the home or are available to support independent living in the home. Informal care was measured by asking participants if they received any help with ADLs or IADLs and who provided that help, e.g. help with ADL and IADL limitations from a spouse/partner, child, relative or other. Participants were also asked if they paid for private non-state care services, namely a personal care attendant or home help. Formal care provided by the State was assessed by asking participants if they were in receipt of community support services, e.g. home help, personal care attendant, meals-on-wheels, and home care packages. Finally, for both informal and formal care, participants were asked how many hours per day and how many days per month they received each of the informal or formal care services. From this data we calculated hours of unpaid and paid informal care per month and hours of formal state provided care per month.

Figures 10 and 11 show the results by CFS categories. There was a progressive increase in the total number of hours of unpaid informal and paid (non-state) care received by participants in the CFS5 to CSF7 groups. There was a 2-fold increase in the total hours of care between those living with mild versus moderate frailty, and a more than 3-fold increase in the total hours of care between those living with mild versus severe frailty (Figure 10). This same pattern was observed when we separated the hours per month of paid non-state care and unpaid informal care (Figure 10). It is notable that participants with mild frailty (CFS5) received informal unpaid but not paid non-state care services. Also of note, is that most non-state care was unpaid informal care provided by family, relatives and others. On a monthly basis, unpaid informal carers provided 100%, 71% and 75% of non-state care hours received by people living with mild, moderate, and severe frailty, respectively.

**Figure 10.**
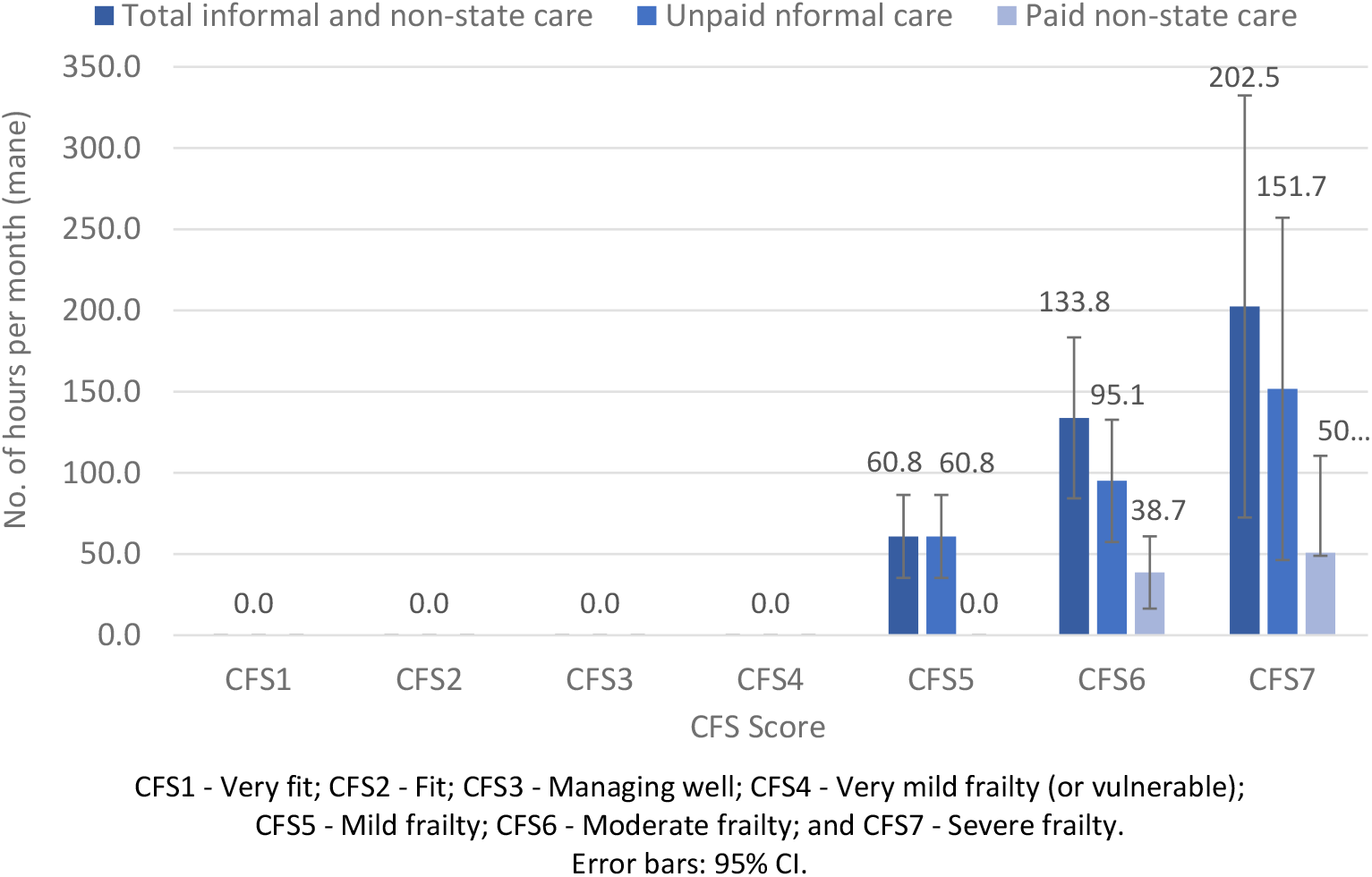
CFS by total hours per month (mean) of unpaid informal and paid non-state care

**Figure 11.**
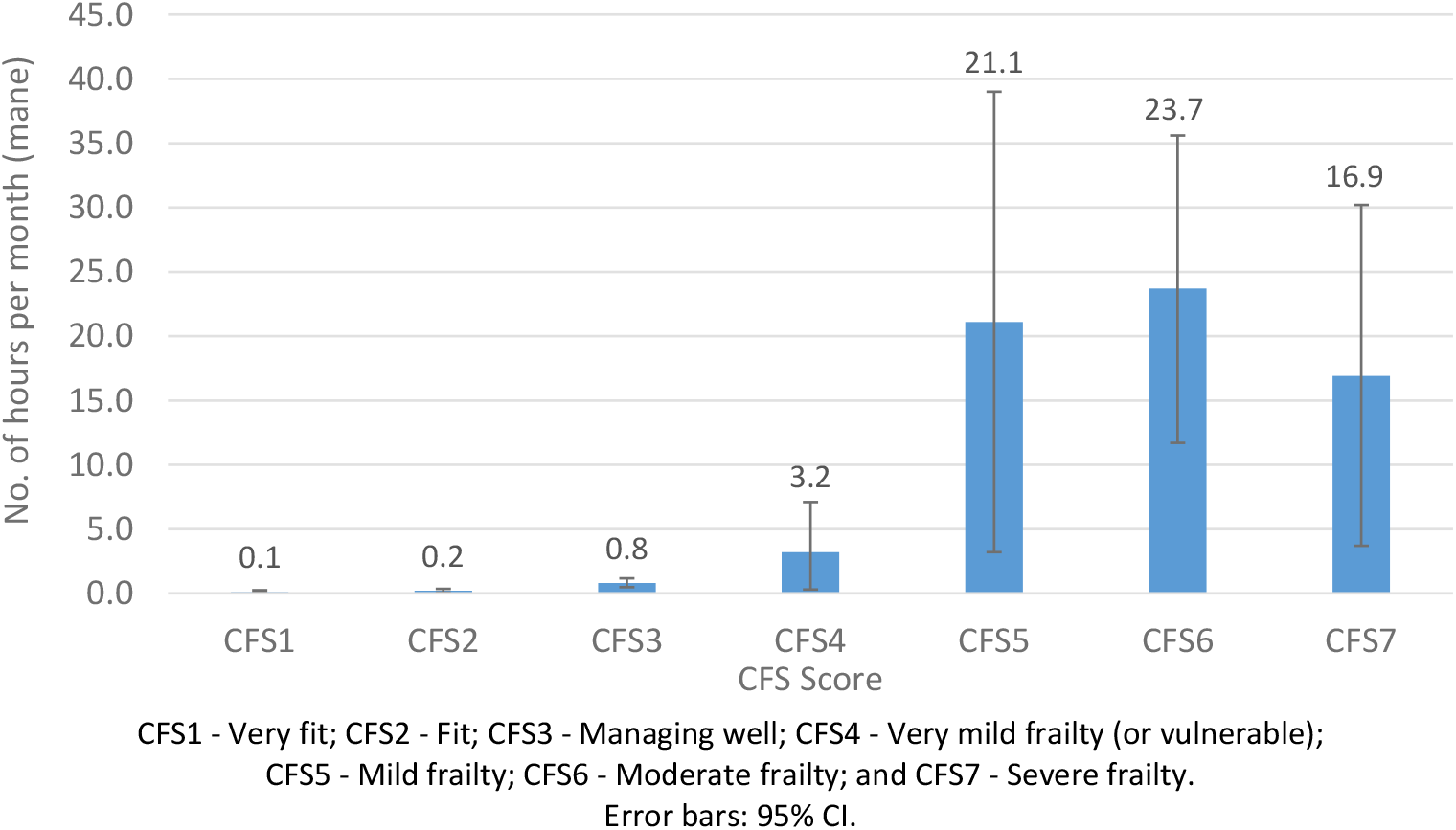
CFS by total hours per month (mean) of formal state care

Total hours per month of formal state care received by CFS categories are provided in Figure 11. There was a progressive increase in the total number of hours of state provided formal care received by participants across groups CFS1 to CSF6 with a possible slight dip in hours received by the CFS7 group. Participants in groups CFS1-4 received ≤3.2 hours per month, while those with mild (CFS5), moderate (CFS6) and severe frailty (CFS7) received 7, 7 and 5-fold more hours of formal state care, respectively. Also of note, is that for every hour of formal state care received per month by those who were mild to severely frail, unpaid informal carers provided between 3-10 hours, and paid non-state carers provided between 0-3 hours of care.

### CFS transitions across Waves 1-5

The alluvial plots showing the transitions between CFS categories and states and deaths at each wave is shown in Figures 12A and 12B, respectively. As expected, the cumulative proportions of deaths and missing data increased across waves. The results of the alluvial plots suggested a dynamic picture of CFS transitions in TILDA over 8 years, both for individual CFS categories and also considering CFS states: ‘fit’ (CFS 1-3), ‘vulnerable’ (CFS 4) and ‘frail’ (CFS >4).

**Figure 12.**
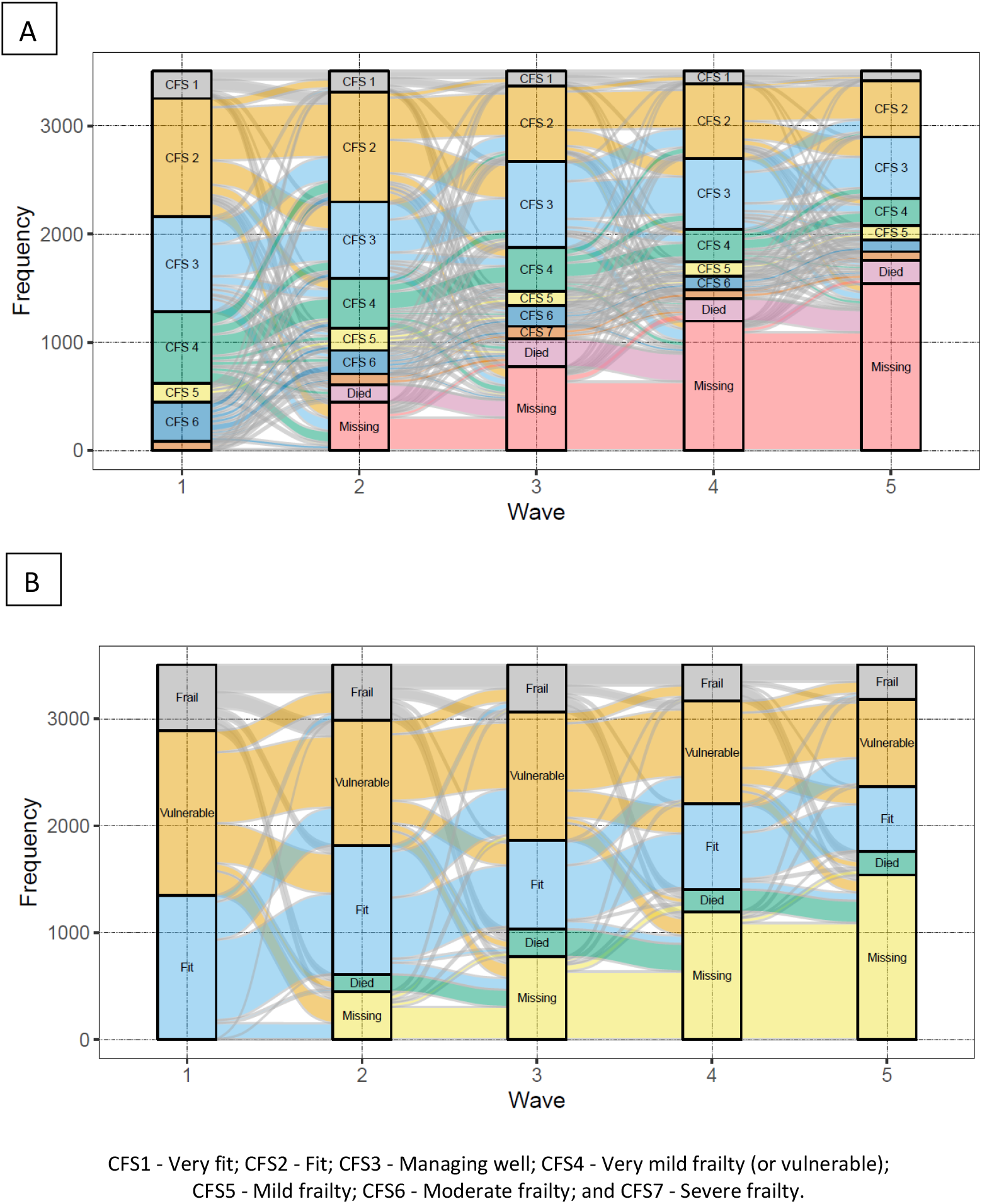
Alluvial plots of CFS group (A) and CFS states (B) transitions in TILDA over 8 years (Waves 1 to 5).

Figure 13 visually shows the results of the multi-state Markov models showing the mean 2-year probability of transitions in CFS states in TILDA. Probabilities from ‘fit’ to ‘vulnerable’, and ‘fit’ to ‘frail’ were 34% and 6%, respectively. ‘Vulnerable’ had a 22% probability of reversal to ‘fit’, and a 16% risk of progression to ‘frail’. ‘Frail’ had a 6% probability of reversal to ‘fit’, a 17% probability of reversal to ‘vulnerable’, and a 25% risk of death. Risks of death for ‘fit’ and ‘vulnerable’ states were low (2% and 5%, respectively). Table 1 shows the transition probabilities with 95% confidence intervals (95% CI).

**Table 1.** Results of the multi-state Markov models showing the mean 2-year probability (with 95% Confidence Intervals) of transitions in CFS states in TILDA.

| CFS State | Fit | Vulnerable | Frail | Died |
| --- | --- | --- | --- | --- |
| Fit | 0.59 (0.54,0.60) | 0.34 (0.31,0.35) | 0.06 (0.05,0.08) | 0.02 (0.01,0.08) |
| Vulnerable | 0.22 (0.21,0.24) | 0.57 (0.55,0.58) | 0.16 (0.14,0.17) | 0.05 (0.04,0.09) |
| Frail | 0.06 (0.06,0.08) | 0.17 (0.15,0.19) | 0.51 (0.49,0.54) | 0.25 (0.23,0.28) |
| Died | 0.00 (0.00,0.00) | 0.00 (0.00,0.00) | 0.00 (0.00,0.00) | 1.00 (1.00,1.00) |

Fit = CFS 1-3; Vulnerable = CFS 4; Frail = CFS 5-7.

**Figure 13.**
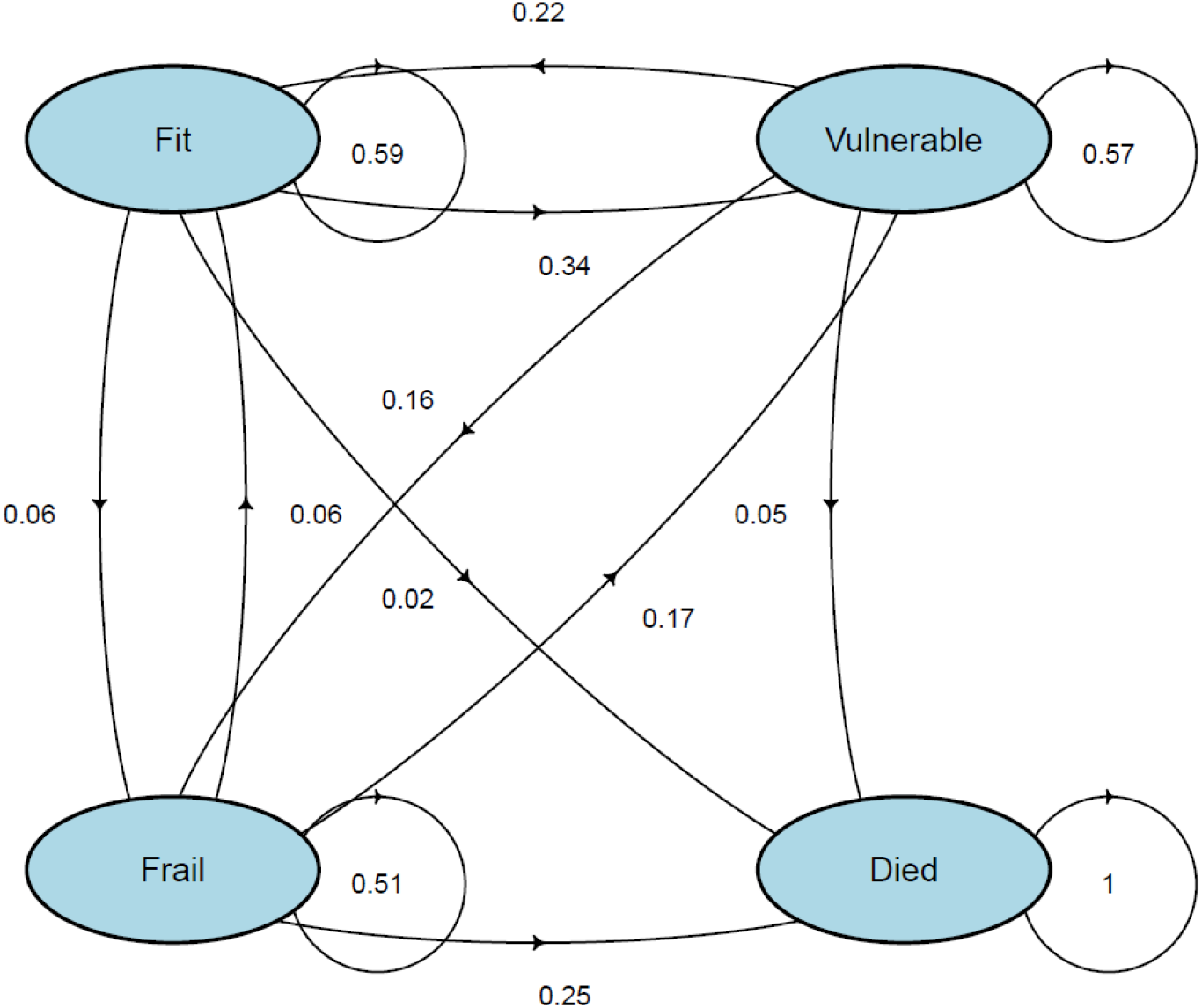
Multi-state Markov models showing the mean 2-year probability of transitions in CFS states.

## Discussion

The aim of the present study was to apply the CFS classification tree to data from adults aged 65 and over from The Irish Longitudinal Study on Ageing (TILDA) and correlate derived CFS categories with patterns of health and social care utilisation in older people in Ireland assessed in Wave 5 of the study (year 2018). In addition, we explored how CFS categories and states changed over 8 years in TILDA between Wave 1 (2010) and Wave 5. Our results suggest that the CFS classification tree was able to stratify the TILDA population aged 65 or more in subgroups with increasing health and social care needs. The CFS classification tree could be used to aid the allocation of health and social care resources in older people in Ireland, but given the frequency of CFS transitions in the population, it is recommended that CFS status is reviewed at least every 2 years in individuals, or sooner when there is a change of individual circumstances.

The National Clinical Programme for Older People has previously highlighted the key roles that the identification of frailty can play in supporting the evolution of age-attuned and age-accommodating services that support timely access to ambulatory care. Indeed, frailty tools could help identify population groups who may have complex care needs as indicated by high levels of health and social care utilisation, for whom timely management of frailty using Comprehensive Geriatric Assessment may help improve both patient and system outcomes (14). Even though no gold standard recommendation exists for the use of the CFS as a frailty identification tool, the CFS has gained currency among professionals for its ease of implementation and ability to incorporate overall clinical judgement in the scoring.

Ageing in place is a key goal of the Irish National Positive Ageing Strategy, which explicitly refers to the Government policy of supporting older people to live in dignity and independence in their own homes and communities for as long as possible. Services such as home helps, home care packages, meals-on-wheels, day centre care and respite care are recognised in the strategy as integral both to supporting this Government’s policy and older people’s own preferred wishes to remain in their own homes (28). Our results suggest scope for considering implementation of the CFS to aid allocation of public resources supporting ageing in place policies.

The results of our study suggest appropriate distributions of CFS categories, as could be expected for a population-based study. As TILDA is a relatively healthy sample of community-dwelling older adults, participants taking part during waves of data collection do not present as very severely frail or terminally ill. Thus, CFS classes 8 and 9 were not observed in the TILDA cohort during data collection waves. Hence, our results cannot be extrapolated to the most vulnerable population of very severely frail and terminally ill individuals, including people living in nursing homes who are often over-represented in those two categories.

In terms of utilisation of medical services, there seemed to be an increase in GP visits between CFS 1 and 6 (3 to 7 visits in the past year), but at CFS 7 it seemed to slightly decline (6), which may correspond to the increase in hospital outpatient visits for the CFS 7 (5 in the past year). CSF 6 and 7 had higher ED attendances and overnight hospital admissions, and a clearly longer hospital length of stay (26-29 days). It is possible that very long hospital length of stays may be indicative of the need to transition to a non-home setting on discharge (e.g. nursing home).

In terms of community care, there were increasing numbers of community services used with increasing CFS levels. Of particular note was the high use of respite and day centre services among those living with frailty in CFS groups 5-7, underscoring the recognition that these services are integral to the Irish National Positive Ageing Strategy, Government’s policy and older people’s own preference to age in place at home. Community rehabilitative services such as OT, physiotherapy, dietician and speech and language therapy services are important to aid functional recovery after hospital discharge following an acute illness, and they may reduce the risk of a new hospital-associated disability becoming a permanent ‘new baseline’ post-discharge. This may also be the case in COVID-19 ‘post-cocooning’ scenarios without a hospital admission. Thus, to avoid having to implement unnecessary and costly large permanent care packages to support home living, it would be prudent to expand the provision of these community and ‘hub’ services beyond the service levels reported here.

With respect to the number of hours received per month of formal state care, paid non-state care and informal unpaid care, all three measures increased steeply between CFS 5 and 7. This reflects the growing burden of ADL and IADL disability with increasing frailty. The vast majority of hours of care per month was provided by unpaid informal carers, i.e. relatives and others, followed by paid non-state care, and formal state provided care. We noted that the CFS5 group received formal and informal care but no paid non-state care, suggesting that while additional care is required this may be balanced against an individual’s ability to pay for additional care. There was a possible reduction in the number of formal care hours per month from CFS6 (23.7) to CFS7 (16.9), which contrasts sharply with the steep increase in informal unpaid and paid non-state care hours between these groups. Again, this suggests increased demand for the provision of formal state community support services for those who are severely frail, the excess of which may currently be carried by unpaid informal carers and by paid non-state provided care.

Our study has various limitations. Firstly, the CFS decision tree is retrospective and does not involve the contemporaneous direct assessment and incorporation of the rater’s clinical judgement on the individual. Indeed, every case is different and individualised decisions should not be replaced by one size fits all. CFS scoring performed by the patient’s side by trained professionals is likely to be more accurate that the retrospective application of a classification tree without clinical judgement being applied. Furthermore, the CFS has only been validated in those aged 65 and over, so we do not recommend use for service planning in those under the age of 65. The TILDA sampling frame does not include people with dementia at baseline or people living in nursing homes, and as such these data may underestimate numbers in receipt of both informal care and formal community support services for the total population aged 65+ years in Ireland.

In conclusion, the CFS classification tree could be used to aid the allocation of health and social care resources in people aged 65+ in Ireland, but given the frequency of CFS transitions in the population, it is recommended that CFS status is reviewed at least every 2 years in individuals, or sooner when there is a change of individual circumstances. Ideally, the scoring of CFS should be part of an expanding, well-trained health and social care workforce with an emphasis on maximisation of community supports, care ‘hubs’ and ambulatory supports.

## Data Availability

TILDA data access information can be accessed on: https://tilda.tcd.ie/data/accessing-data/. Data utilised in this study can be made accessible on reasonable request.

## Appendices

### Appendix 1: Number of chronic conditions: 28 cardiovascular and chronic conditions

1. High blood pressure or hypertension
2. Angina
3. A heart attack (including myocardial infarction or coronary thrombosis)
4. Congestive heart failure
5. Diabetes or high blood sugar
6. A stroke (cerebral vascular disease)
7. Ministroke or TIA
8. High cholesterol
9. A heart murmur
10. An abnormal heart rhythm (including Atrial Fibrillation)
11. Any other heart trouble (please specify)
12. Chronic lung disease such as chronic bronchitis or emphysema
13. Asthma
14. Arthritis (including osteoarthritis, or rheumatism)
15. Osteoporosis, sometimes called thin or brittle bones
16. Cancer or a malignant tumour (including leukaemia or lymphoma but excluding minor skin cancers)
17. Parkinson’s disease
18. Any emotional, nervous or psychiatric problems such as depression or anxiety
19. Alcohol abuse or substance abuse
20. Alzheimer’s disease
21. Dementia, organic brain syndrome, senility
22. Serious memory impairment
23. Stomach ulcers
24. Varicose ulcers (an ulcer due to varicose veins)
25. Cirrhosis, or serious liver damage
26. Cataracts
27. Glaucoma
28. Age related macular degeneration

## Notes

### Competing Interest Statement

The authors have declared no competing interest.

